# Proteomic and Metabolomic Characterization of COVID-19 Patient Sera

**DOI:** 10.1101/2020.04.07.20054585

**Authors:** Bo Shen, Xiao Yi, Yaoting Sun, Xiaojie Bi, Juping Du, Chao Zhang, Sheng Quan, Fangfei Zhang, Rui Sun, Liujia Qian, Weigang Ge, Wei Liu, Shuang Liang, Hao Chen, Ying Zhang, Jun Li, Jiaqin Xu, Zebao He, Baofu Chen, Jing Wang, Haixi Yan, Yufen Zheng, Donglian Wang, Jiansheng Zhu, Ziqing Kong, Zhouyang Kang, Xiao Liang, Xuan Ding, Guan Ruan, Nan Xiang, Xue Cai, Huanhuan Gao, Lu Li, Sainan Li, Qi Xiao, Tian Lu, Yi Zhu, Huafen Liu, Haixiao Chen, Tiannan Guo

## Abstract

Severe COVID-19 patients account for most of the mortality of this disease. Early detection and effective treatment of severe patients remain major challenges. Here, we performed proteomic and metabolomic profiling of sera from 46 COVID-19 and 53 control individuals. We then trained a machine learning model using proteomic and metabolomic measurements from a training cohort of 18 non-severe and 13 severe patients. The model correctly classified severe patients with an accuracy of 93.5%, and was further validated using ten independent patients, seven of which were correctly classified. We identified molecular changes in the sera of COVID-19 patients implicating dysregulation of macrophage, platelet degranulation and complement system pathways, and massive metabolic suppression. This study shows that it is possible to predict progression to severe COVID-19 disease using serum protein and metabolite biomarkers. Our data also uncovered molecular pathophysiology of COVID-19 with potential for developing anti-viral therapies.

## INTRODUCTION

Coronavirus disease 2019 (COVID-19) is an unprecedented global threat caused by severe acute respiratory syndrome coronavirus 2 (SARS-CoV-2). It is currently spreading around the world rapidly. The sudden outbreak and accelerated spreading of SARS-CoV-2 infection have caused substantial public concerns. Within about two months, close to one million individuals worldwide have been infected, leading to about 50,000 deaths. At the time of writing this manuscript, about 100,000 new infections are reported daily.

Most COVID-19 studies have focused on its epidemiological and clinical characteristics(Ghinai et al., 2020; Guan et al., 2020; Li et al., 2020b; Team, 2020). About 80% of patients infected with SARS-CoV-2 displayed mild symptoms with good prognosis(Team, 2020). They usually recover with, or even without, conventional medical treatment, and therefore are classified as mild or moderate COVID-19 (Medicine, 2020; Thevarajan et al., 2020). However, about 20% of patients suffer from respiratory distress and require immediate oxygen therapy or other inpatient interventions, including mechanical ventilation (Medicine, 2020; Murthy et al., 2020; Wu and McGoogan, 2020). These patients, classified as clinically severe or critical life-threatening infections, are mainly diagnosed empirically based on a set of clinical characteristics, such as respiratory rate (≥ 30 times/min), mean oxygen saturation (≤ 93% in the resting state) or arterial blood oxygen partial pressure/oxygen concentration (≤ 300 mmHg) (Medicine, 2020). However, patients exhibiting these clinical manifestations have already progressed to a clinically severe phase and require immediate access to specialized intensive care; otherwise, they may die rapidly. Therefore, it is critical to develop new approaches to predict early which cases will likely become clinically severe. In addition, effective therapy for severe patients remains speculative, largely due to limited understanding of SARS-CoV-2 pathogenesis.

In this study, we hypothesized that SARS-CoV-2 induces characteristic molecular changes that can be detected in the sera of severe patients. These molecular changes may shed light on therapy development for COVID-19 patients. To test this hypothesis, we applied cutting-edge proteomic (Aebersold and Mann, 2016; Li et al., 2020a) and metabolomic (Hou et al., 2020; Lee et al., 2019) technologies to analyze the proteome and metabolome of sera from severe COVID-19 patients and several control groups.

## RESULTS

### Proteomic and metabolomic profiling of COVID-19 sera

We procured a cohort of patients (Zheng et al., 2020) containing 21 severe COVID-19 patients, in which 11 sera from severe patients were collected one to six days before the patients were clinically assessed as severe. Sera from the remaining eight patients were collected within about 3 days after diagnosed as severe cases (Figure 1A, Table 1, Table S1). Controls with matched epidemiological features were included to identify severity-related molecular alterations. These controls were 28 healthy subjects, 25 non-COVID-19 patients (negative for the SARS-CoV-2 nucleic acid test) with similar clinical characteristics as COVID-19 patients, and 25 non-severe COVID-19 patients. A serum sample was obtained from each patient within a few days after hospital admission, with a few exceptions when samplings were performed at later disease stages.

**Table 1.**
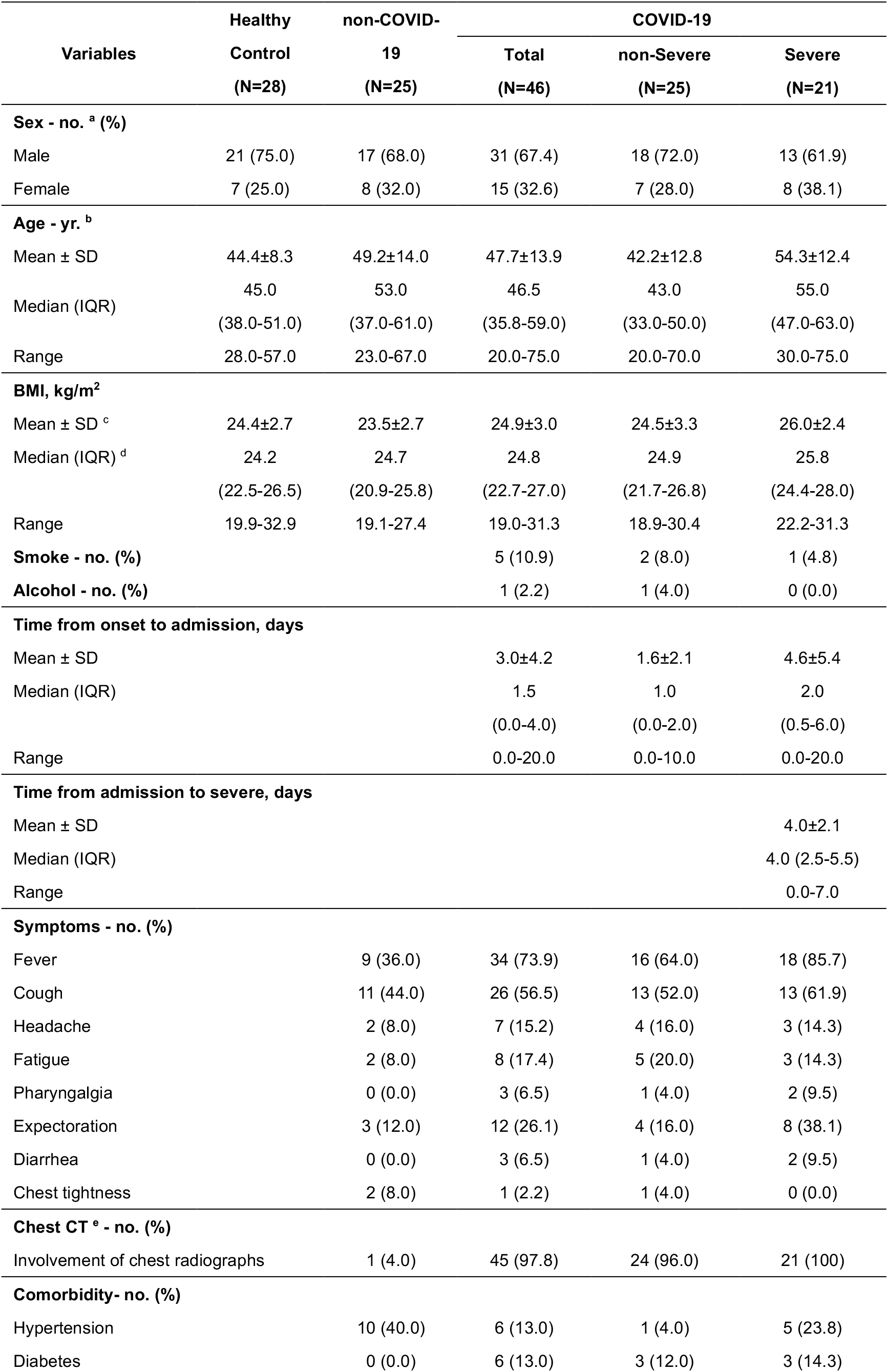
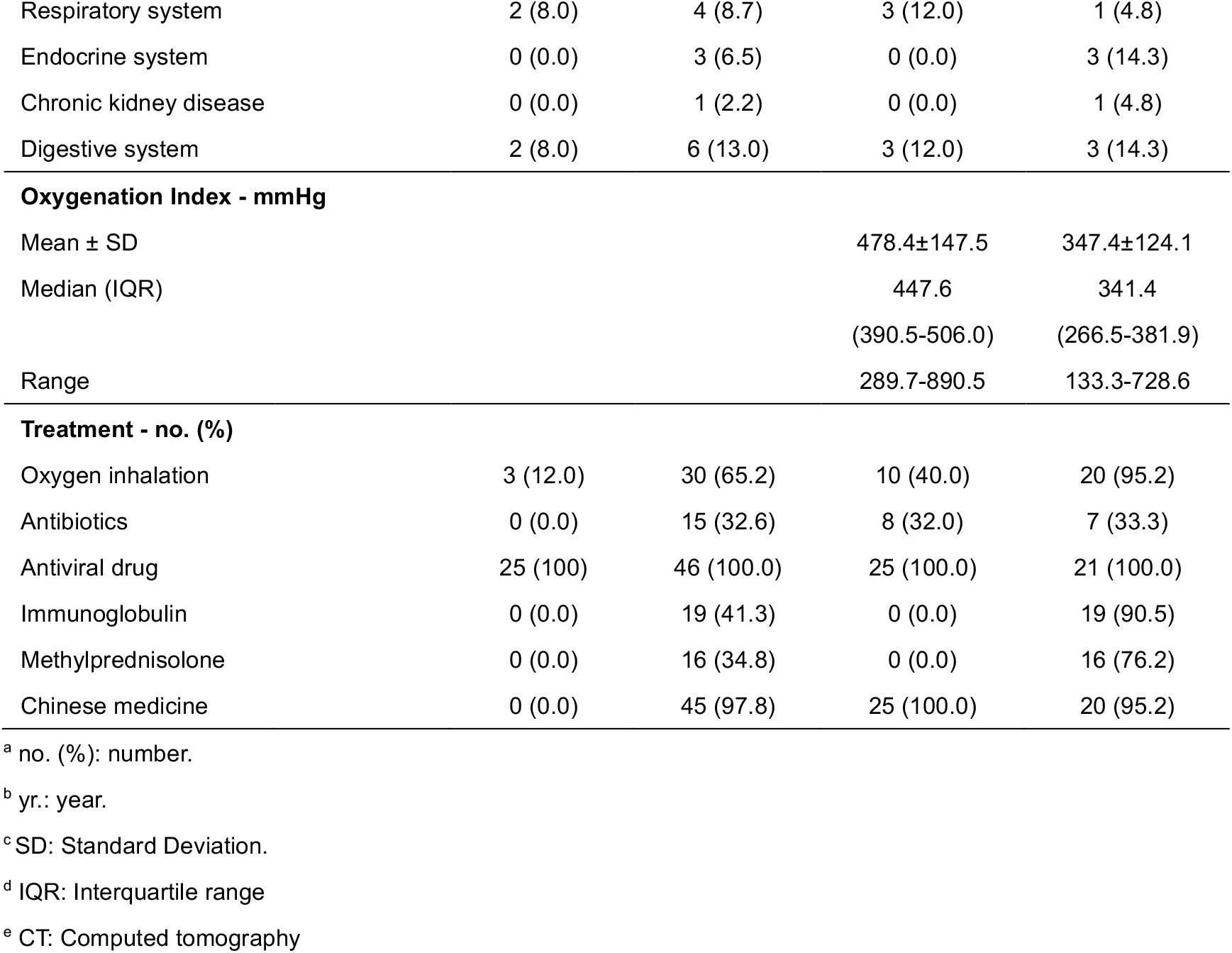
Demographics and baseline characteristics of COVID-19 patients.

**Figure 1.**
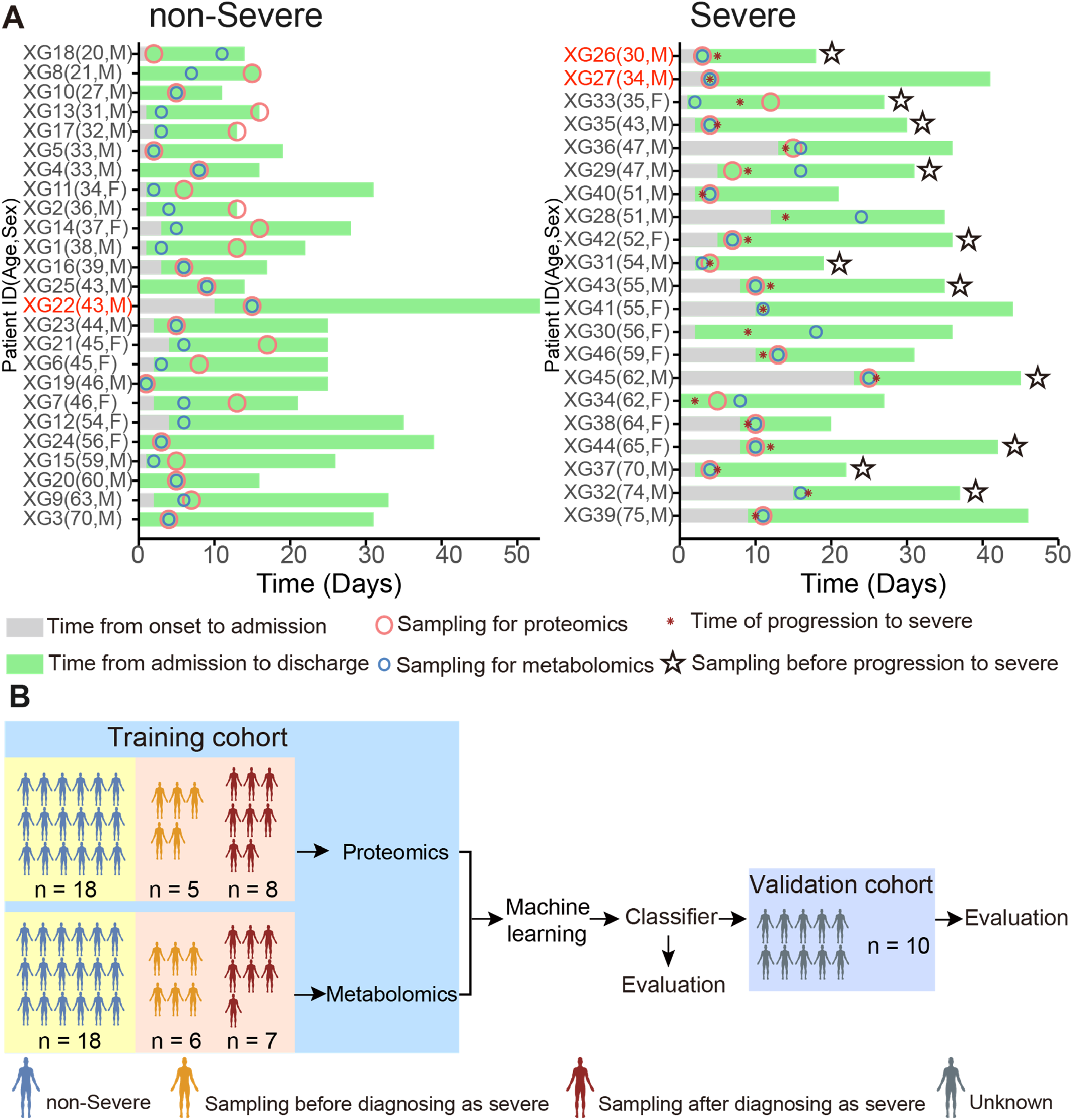
Summary of COVID-19 patients and machine learning design. Summary of COVID-19 patients, including non-severe (n=25) and severe (n=21) patients with more details in Table S1. Patients labeled in red (Y-axis) indicate chronic infection of viruses including HBV. (B) Study design for machine learning-based classifier development for severe COVID-19 patients. We first procured samples in a training cohort for proteomic and metabolomic analysis. The classifier was then validated in an independent cohort.

We used stable isotope labeled proteomics strategy TMTpro (16plex) (Li et al., 2020a) and UPLC-MS/MS untargeted metabolomics approach to analyze 92 undepleted sera from 86 individuals. Altogether 894 proteins were quantified and 941 (including 36 drugs and their metabolites) metabolites were identified with authentic compound library searching. For metabolomic analysis, both hydrophilic and hydrophobic molecules were analyzed using both positive and negative ionization to cover various endogenous biochemical classes. Our data were acquired with high degree of consistency and reproducibility. In quality control analysis, the median coefficient of variance (CV) values for proteomic and metabolomic data were 10% and 5%, respectively (Figure S1A). Remarkably, sera from SARS-CoV-2 infected patients were well resolved from healthy individuals, and the sera from severe patients displayed distinct proteomic and metabolomic patterns compared to those from other groups (Figures S1B-C).

### Identification of severe patients using machine learning

We next investigated the possibility of classify the severe COVID-19 patients based on the molecular signatures of proteins and metabolites (Table S2). We built a random forest machine learning model based on proteomic and metabolomic data from 18 non-severe and 13 severe patients (Figure 1B), leading to prioritization of 29 important variables including 22 proteins and 7 metabolites (Figure 2A-B). This model reached an AUC of 0.957 in the training set (Figure 2C). One non-severe patient, XG3, was incorrectly classified as severe (Figure 2D), possibly because this 70-year-old male patient was the oldest individual in this cohort (Figure 1A). For patient XG40, the reason of incorrectly classified is unclear.

**Figure 2.**
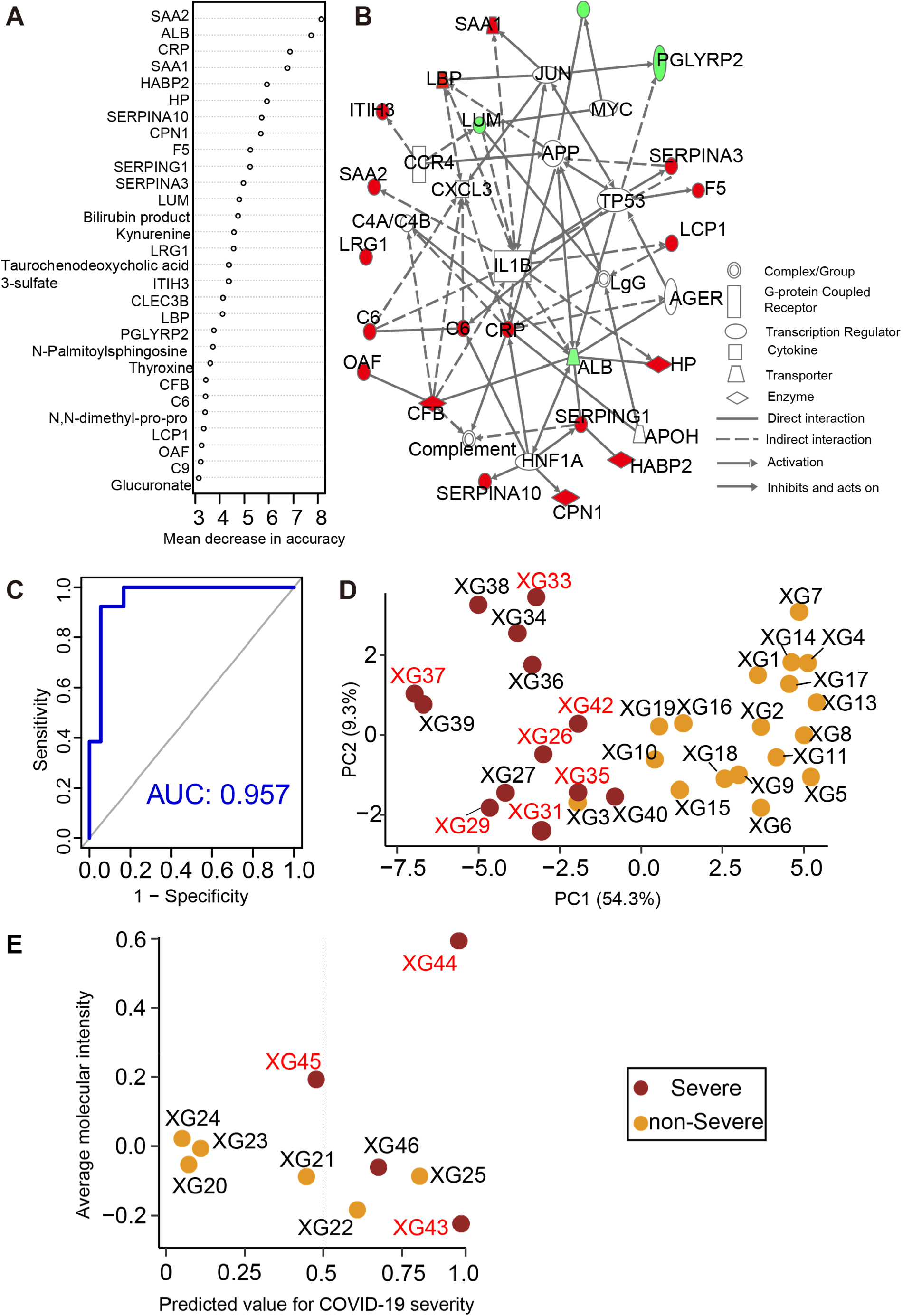
Separation of severe and non-severe COVID-19 patients by machine learning of proteomic and metabolomic features. (A) Top 22 proteins and 7 metabolites prioritized by random forest analysis ranked by the mean decrease in accuracy. (B) Network of prioritized proteins appeared in the classifier. (C) ROC of the random forest model in the training cohort. (D) Principal Components Analysis (PCA) plot of COVID-19 patients from the training cohort. (E) Performance of the model in the validation cohort of ten COVID-19 patients. Patients labeled in red received serum test before they were diagnosed as severe.

We then tested the model on an independent cohort of ten patients (Figure 2E, Table S3). All severe patients were correctly identified, except one patient, XG45, a 62-year-old male who had received traditional Chinese medicines during more than 20 days before admission. This individual was the patient with the longest pre-admission treatment in this cohort and the administration of traditional Chinese medicines might have also accounted for the incorrect prediction. Another incorrectly classified non-severe patient was XG22, a 43 year-old male with chronic HBV infection and diabetes who had the longest hospitalization (>50 days) among all non-severe patients. Indeed, our molecular-based prediction flagged him as an outlier, suggesting that his prolonged treatment history might have interfered with the expression of the panel of the 29 variables. For reasons yet unclear, XG25, a 43-year-old male non-severe case, was incorrectly classified as severe.

### Proteomic and metabolomic changes in severe COVID-19 sera

We found that 105 proteins were differentially expressed in COVID-19 patients, but not in the non-COVID-19 patients (Figure S2, S4). After correlating their expression with clinical disease severity (Figure S5), 93 proteins showed specific modulation in severe patients. Pathway analysis and network enrichment analyses of the 93 differentially expressed proteins revealed three pathways (Figure S6), namely activation of the complement system, macrophage function and platelet degranulation, involving 50 proteins (Figure 3A) in the sera of severe patients. Similarity, 373 and 204 metabolites were found with differential abundance in COVID-19 patients and continually changed correlated with disease severity in our metabolomics data, respectively (Figure S3, S4). Correspondingly, 82 metabolites were involved in the three biological processes. We summarize the key dysregulated molecules in Figure 5 and discussed in the following sections.

**Figure 3.**
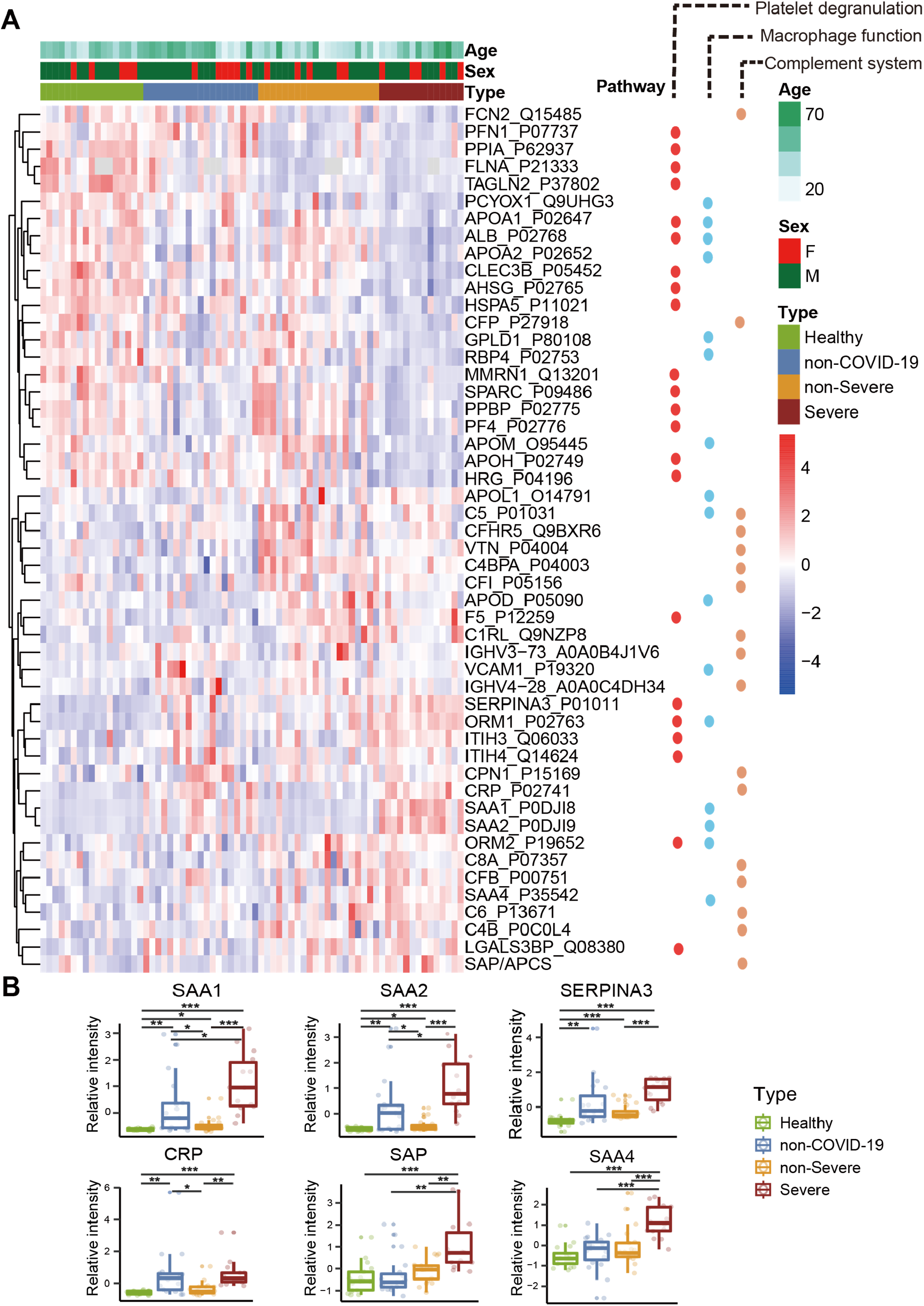
Dysregulated proteins in COVID-19 sera. (A) Heatmap of 50 selected proteins whose regulation concentrated on three enriched pathways. The expression level change (z-scored original value) of six selected proteins of interest with significance indicated by the asterisks (unpaired two sided Welch’s t test. p value: ***, <0.001; **, <0.01; *, <0.05)

**Figure 5.**
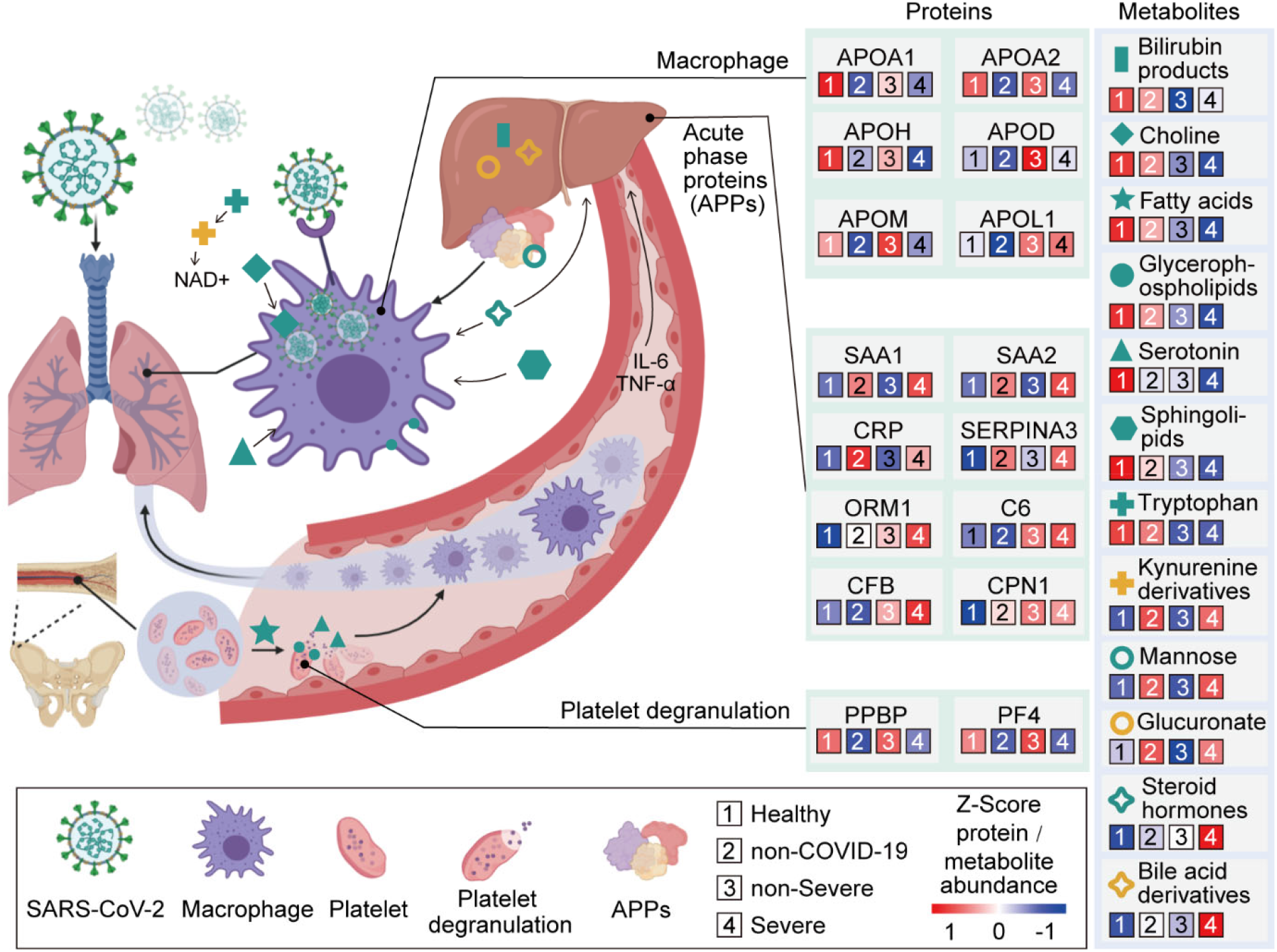
Key proteins and metabolites characterized in severe COVID-19 patients in a working model. SARS-CoV-2 may target alveolar macrophages via ACE2 receptor with increasing level of cytokines including IL-6 and TNF-α, which subsequently induce the elevation of various APPs such as SAP, CRP, SAA1, SAA2 and C6 which are significantly upregulated in the severe group. Proteins involved in macrophage, lipid metabolism and platelet degranulation were indicated with their corresponding expression levels in four patient groups.

### Dysregulated macrophage and lipid metabolism

Our data uncovered dysregulation of multiple apolipoproteins including APOA1, APOA2, APOH, APOL1, APOD and APOM (Figure 3A). Most of them are associated with macrophage function and were down-regulated. Decrease of APOA1 in serum has been reported during the transition of COVID-19 patients from mild to severe illness (Nie et al., 2020). The APOM in serum of severe patients was downregulated compared with healthy and non-severe COVID-19 patients. Regulation of serum APOM has also been observed in hepatitis B virus (HBV) patients (Gu et al., 2011).

Consistent with these proteomic findings, we also detected dysregulated metabolites involved in lipid metabolism. Accumulation of 16 steroid hormones in COVID-19 patients may contribute to macrophage modulation. Steroid hormones, including progesterone, androgens, estrogens and bile acids can promote the activity of macrophages (Vernon-Roberts, 1969). Specifically, glucocorticoids were recently reported to be clinically efficacious (Wang et al., 2020b). We detected increased 21-hydroxypregnenolone, the essential intermediate for synthesizing corticosterone, suggesting that corticosterone biosynthesis may be a protective mechanism against SARS-CoV-2 infection.

We also found evidence of significant activation of the kynurenine pathway. Metabolites of kynurenate, kynurenine, 8-methoxykynurenate were enriched in COVID-19 patients. Nicotinamide adenine dinucleotide (NAD+), the cofactor in many cellular redox reactions, can be synthesized from tryptophan by the kynurenine pathway and operates as a switch for macrophage effector responses (Minhas et al., 2019).

The macrophage process is closely related to lipid metabolism. Over 100 lipids were down-regualted in severe patients. Our data showed decreased sphingolipids in both non-severe and severe COVID-19 patients (Figure 4A). Sphingolipids and glycerophospholipids are important components of biomembranes, which mediate signal transduction and immune activation processes. Sphingolipids, such as sphingosine-1-phosphate, can induce macrophage activation, inhibit macrophage apoptosis, and promote migration of macrophages to inflammatory sites (Weigert et al., 2009). Phagocytosis and platelet degranulation are coupled with changes in biomembrane lipid composition and fluidity, and modulate the production of glycerophospholipids (Rouzer et al., 2007). In this study, we found continuous decrease of glycerophospholipids after SARS-CoV-2 infection. Glycerophospholipids and fatty acids such as arachidonic acid have been found significantly elevated in HCoV-229E-infected cells, and exogenous supplement of arachidonic acid significantly suppressed HCoV-229E and MERS-CoV replication (Yan et al., 2019). Our data suggest severe COVID-19 patients might benefit from this therapeutical strategy as well.

**Figure 4.**
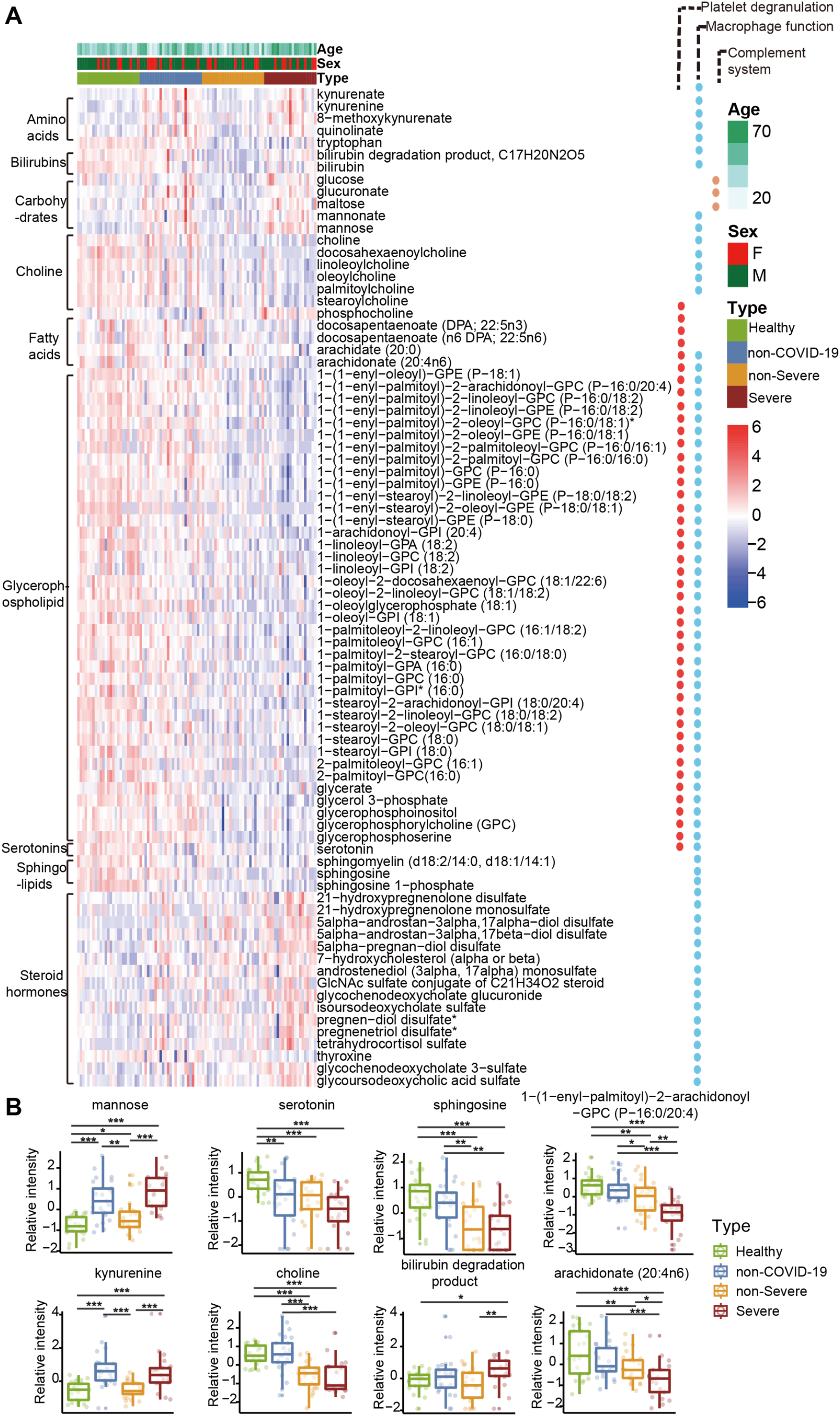
Dysregulated metabolites in COVID-19 sera. (A) Heatmap of 82 regulated metabolites belonging to six major classes: fatty acids, steroids, glycerophospholipid, sphingolipid, choline and serotonins. (B) The expression level change (z-scored log2 original value) of six regulated metabolites from each metabolite class with significance indicated by the asterisks as described in Figure 3.

Choline and its derivatives were down-regulated in COVID-19 patients, particularly in severe cases; phosphocholine, the intermediate product for producing phosphatidylcholine (PC) was up-regulated (Figure 4B). This was probably due to activated macrophage-mediated immunity (Sanchez-Lopez et al., 2019). Polarization of macrophages in response to pathogens requires increased absorption of choline for PC formation, thereby promoting cytokine secretion (Sanchez-Lopez et al., 2019).

### Activated acute phase proteins (APPs) and the complement system

We detected 10 APPs among 20 proteins that are differentially expressed between non-severe and severe groups (Figure 2A). They are involved at the early stages of immune responses to virus infection. Among the most significantly upregulated in severe sera were APPs, including serum amyloid A-1 (SAA1), serum amyloid A-2 (SAA2), serum amyloid A-4 (SAA4), C-reactive protein (CRP), alpha-1-antichymotrypsin (SERPINA3) and serum amyloid P-component (SAP/APCS) (Figure 3B). Some of them are known to be biomarkers for viral infections, including SAA1, SAA2 and CRP. While CRP has been associated with COVID-19, the other proteins have not previously been reported in COVID-19 (Liang et al., 2020). SAA1 was reported to be elevated in severe SARS patients, but was not specific to SARC-CoV (Pang et al., 2006). As a major contributor to acute phase response, complement system plays a crucial role in eliminating invading pathogens in the early stage of infection. Among those APPs, two proteins belong to the complement membrane attack complex, including complement 6 (C6) and complement factor B (CFB). Two other proteins, Properdin (CFP) and Carboxypeptidase N catalytic chain (CPN1), are regulators of complement system (Figure 3).

We also observed an accumulation of mannose and its derivatives in severe patients. In the complement system, binding of mannose to lectin leads to cleaveage of C2 and C4, which then form a C3 convertase to promote complement activation (Ricklin et al., 2010).

### Suppressed platelet degranulation in severe sera

Fifteen of 17 proteins involved in platelet degranulation were down-regulated in SARS-CoV-2 infected patients, which may be associated with observed thrombocytopenia in this patient cohort (Zheng et al., 2020). Low platelet count is also reported to be associated with severe COVID-19 and mortality (Lippi et al., 2020). Two of the most intriguing proteins down-regulated in severe patients are platlet expressing chemokines pro-platelet basic protein (PPBP; also called macrophage-derived growth factor) and platelet factor 4 (PF4). PF-4 was identified as a broad-spectrum HIV-1 inhibitor at the level of virus attachment and entry via interaction with the major viral envelope glycoprotein gp120 (Auerbach et al., 2012). In another sera proteomic investigation of SARS, they found decreasing PF4 was associated with poor prognosis (Poon et al., 2012), in consistent with our findings regarding COVID-19.

Most enterochromaffin cell-derived serotonins (5-hydroxytryptamine [5-HT]) are transported to platelets for storage and release (Baganz and Blakely, 2013). Our previous data showed that when platelet counts in COVID-19 patients decrease as the severity of the disease increase (Zheng et al., 2020), serotonin (Figure 4B) level decreases accordingly. Compared with the healthy group, serotonin in non-severe and severe COVID-19 patients decreased by 2.07-fold (p = 1.86e-04) and 3.31-fold (p = 9.07e-07), respectively.

We also detected low levels of fatty acids such as arachidonate and docosapentaenoate in COVID-19 patients (Figure 4), which may be related to their decreased platelet counts. Fatty acids including arachidonate (20:4n6) are active factors of platelet degranulation. A study on H7N9 reported that H7N9 infection led to suppression of various fatty acids including palmitic acid (Sun et al., 2018). We also found palmitic acid decreased in severe COVID-19 patients.

### Massive suppression of amino acids in the sera of COVID-19 patients

More than 100 decreased metabolites in the sera of COVID-19 patients are amino acids and their derivatives were found significantly decreased in the sera of COVID-19 patients compared to their levels in the healthy controls, while their levels were either unchanged or even increased in the sera of non-COVID-19 patients. Enriched in these metabolites are ten metabolites involved in arginine metabolism including glutamate, arginine, N-(l-arginino)succinate, citrulline, ornithine, glutamine, 2-oxoglutarate, N-acetyl-L-glutamate, urea, and fumarate. In addition, some arginine derivatives such as argininate, asymmetric dimethylarginine, symmetric dimethylarginine, homoarginine and N-acetylarginine, were also significantly decreased in the sera of non-severe COVID-19 patients. It has been reported that arginine metabolism is suppressed in severe fever with thrombocytopenia syndrome caused by a SFTS bunyavirus (SFTSV)(Li et al., 2018b). Decreased arginine levels in SFTSV patients was associated with impaired anti-SFTSV functions of T cells.

## DISCUSSION

### Prediction of severe COVID-19 patients

Although COVID-19 can be diagnosed effectively by nucleic acid-based methods at an early stage, it is equally critical to identify severe COVID-19 patients before their manifestion of severe symptoms to minimize mortality. In this study we show that severe cases can be predicted by molecular signatures of metabolites and proteins using a machine learning model based on the expression levels of 22 serum proteins and seven metabolites (Figure 2A-B). We achieved an overall accuracy of 93.5% in the training set. Prediction of two patients did not match clinical diagnosis. One of them is non-severe individual who was the oldest patient in the training cohort. Remarkably, nine severe patients were correctly identified retrospectively based on the analysis of their sera collected one to six days before they were clinically diagnosed as having deteriorated to a severe state (Figure 1, Figure 2D), suggesting that their sera protein and metabolite signatures at the sampling time were already pointing to further deterioration into severe state even when severe clinical symptoms have not started to appear yet.

The proteins and metabolites used in our classifier (Figure 2A) contain several known biomarkers for viral infections, such as SAA2, SSA1 and CRP, which have already been used empirically to monitor the severity of COVID-19. Our study suggests that more characteristic molecular changes at protein and metabolite levels can be used to build a predictive model for the prospective identification of severe cases. The classifier also included exceptionally high levels of other acute phase proteins, including SERPINA3, among others (Figure 2A-B). Our data suggest potential benefits of broader testing of these proteins in newly diagnosed cases to identify which COVID-19 patients are likely to progress to severe disease. The model contains molecules involved in hepatic damage. The elevation of glucose, glucuronate, bilirubin degradation product and four bile acid derivatives, suggests suppressed hepatic detoxification (Rowland et al., 2013). Vascular cell adhesion protein 1 (VCAM-1) which helps to regulate transendothelial migration of leukocytes by stimulating production of reactive oxygen species (ROS), was upregulated in our data. As apotent antioxidant and inhibitor of VCAM-1 dependent cellular events (Keshavan et al., 2005), bilirubin was found to be down-regulated in our metabolomic data.

Seven patients were correctly classified in the independent validation cohort containing ten patients. Two of them could be explained by the patients’ complex comorbidity and medication history. The relatively small sample size necessitates future validation studies in independent cohorts.

### Molecular insights for pathogenesis of SARS-CoV-2 infection

Our data shed light on the molecular changes reflected in COVID-19 sera which could potentially yield critical diagnostic markers or therapeutic targets for managing severe COVID-19 patients (Figure 5). These molecular derangements may originate from binding of SARS-CoV-2 to alveolar macrophages via the ACE2 receptor (Hoffmann et al., 2020), resulting in release of IL-6 and TNF-α (Mehta et al., 2020) by macrophages (Gabay and Kushner, 1999). In response to elevated cytokines, especially IL-6 which triggers fever, the liver releases various APPs. Activation of APPs is accompanied with the immunogenesis or organic damage (Gabay and Kushner, 1999). Our metabolomics data also provide plausible evidence for hepatic injury. In physiological condition, hormone or bilirubin binds to glucuronate, a derivative of glucose, for liver detoxification (Rowland et al., 2013). The elevation of glucose, glucuronate, bilirubin degradation product and four bile acid derivatives in severe patients, indicating the potential declined detoxification function (Figures 4-5). Indeed, our data showed an upregulation of multiple APPs, including CRP and major attack complexes (MACs) in the severe sera. CRP can activate the complement system (Chirco and Potempa, 2018). This on the one hand leads to enhanced cytokine and chemokine production, potentially contributing to “cytokine storm”; and on the other overly recruits macrophages from the peripheral blood, which could result in acute lung injury(Chirco and Potempa, 2018; Narasaraju et al., 2011). Because about 50% of platelets are produced in the lung (Lefrancais et al., 2017), platelet may in turn respond to lung injury and activate macrophases by degranulation (Mantovani and Garlanda, 2013), which may further add to “cytokine storm”. A recent necropsy report revealed alveolar macrophage infiltration and activation in severe COVID-19 patients (Liao M., 2020), supporting our findings.

### Insights for COVID-19 therapeutics

To date, few other therapies are proven effective for severe COVID-19 patients. Most of patients receive standard supportive care and antiviral therapy (Wang et al., 2020a). Corticosteroid treatment which are effectively in suppressing MERS-CoV and SARS-CoV (Arabi et al., 2018), but showed negligible effect on COVID-19 patients and may even have induced lung injury (Russell et al., 2020). The molecular changes revealed in this study in the COVID-19 sera have allowed us to propose potential new therapeutic strategies for the severe patients.

Our proteomic data showed that proteins related to platelet degranulation were substantially down-regulated in severe patients, a finding which was confirmed by low platelet counts (Zheng et al., 2020). The association between thrombocytopenia and viral infection has been observed in SARS-CoV (Zou et al., 2004), hepatitis C virus (HCV) (Assinger, 2014), and Dengue virus (Wilder-Smith et al., 2004). We thus recommend watching closely monitoring changes in platelets and making interventions, such as infusions of thrombopoietin (TPO) as necessary, particularly when liver or kidney injuries occur.

Complement activation suppresses virus invasion, and may lead to inflammatory syndromes (Barnum, 2016). Our data showed a general up-regulation of complement system proteins, including MAC proteins such as C5, C6 and C8. Suppression of complement system has been reported as an effective immunotherapeutics in SARS-infected mouse model (Gralinski et al., 2018). C5a has been reported as highly expressed in severe SARS and MERS patients as well (Wang et al., 2015). Inhibition of C5a has been reported to alleviate viral infection-induced acute lung injury (Garcia et al., 2013; Jiang et al., 2018; Sun et al., 2014). Our data suggest that severe COVID-19 patients might benefit from suppression of complement system.

The coronavirus are enveloped, positive strand RNA viruses, the replication and assembly of which consume large amount of lipids. Our metabolomics results showed that more than 100 lipids including glycerophospholipid, sphingolipids and fatty acids were down-regulated in COVID-19 patients sera, probably due to rapid replication of the virus. Glycerophospholipid, sphingolipids (one of the components of lipid rafts) and fatty acids have been reported to play an important role in the early development of enveloped viruses (Schoggins and Randall, 2013). Suppression of cholesterol synthesis by MβCD has been reported inhibiting the production of SARS-CoV particles released from infected Vero E6 cells (Li et al., 2007). Drugs inhibiting lipid synthesis such as statin have been proposed to treat HCV (Heaton and Randall, 2011) and COVID-19 (Fedson et al., 2020). Our data also supports this potential therapeutics for severe COVID-19 patients.

SARS-CoV-2 is highly infectious, applying huge pressure to the medical system worldwide. Upon COVID-19 outbreak, limited information of this pathogen was available, which restricted the collection of clinical specimens for this study. The reason for the small sample size of this study were biosafety constraints.

In conclusion, this study presents a systematic proteomic and metabolimic investigation of sera from multiple COVID-19 patient groups and control groups. We show that feasibility of predicting severe COVID-19 patients based on a panel of serum proteins and metabolites. Our data offer a landscape of blood molecular changes induced by SARS-CoV-2 infection, which may provide useful diagnositic markers and therapeutic targets.

## Data Availability

All data are available in the manuscript or the supplementary material. The proteomics and metabolomics data are deposited in ProteomeXchange Consortium (https://www.iprox.org/). Project ID: IPX0002106000. All the data will be publicly released upon publication.

## ACKNOWLEDGEMENTS

This work is supported by grants from Westlake Special Program for COVID-19 (2020), and Tencent foundation (2020), National Natural Science Foundation of China (81972492, 21904107, 81672086), Zhejiang Provincial Natural Science Foundation for Distinguished Young Scholars (LR19C050001), Hangzhou Agriculture and Society Advancement Program (20190101A04). We thank Drs. R. Aeberold, O.L. Kon, H. Yu, D. Li, and the Guomics team for invaluable comments to this study. We thank Westlake University Supercomputer Center for assistance in data storage and computation.

## AUTHOR CONTRIBUTIONS

T.G., H.C., H.L., B.S. and Y.Z. designed and supervised the project. B.S., X.B., J.D., Y.Z., J.L., J.X., Z.H., B.C., J.W., H.Y., Y.Z., D.W. and J.Z. collected the samples and clinical data. X.Y., Y.S., F.Z., R.S., L.Q., W.G., W.L., S.L., H.C., X.L., X.D., G.R., N.X., X.C., H.G., L.L., S.L., Q.X. and T.L. conducted proteomic analysis. Data were interpreted and presented by all co-authors. C.Z., S.Q., Z.K. and Z.K. conducted metabolomic analysis. T.G. wrote the manuscript with inputs from co-authors.

## MATERIALS AND METHODS

### Patients and samples

Our team procured serum samples from 46 patients who visited Taizhou Public Health Medical Center during January 23 and February 4, 2020. They were diagnosed as COVID-19 according to the Chinese Government Diagnosis and Treatment Guideline (Trial 5th version)(Medicine, 2020). For diagnosing COVID-19, nucleic acid from sputum or throat swab was extracted using nucleic acid extractor (Shanghai Zhijiang, China, EX3600) and virus nucleic acid extraction reagent (Shanghai Zhijiang, China, NO. P20200201) was used to extract nucleic acid. Fluorescence quantitative PCR (ABI7500) and SARS-CoV-2 nucleic acid detection kit (triple fluorescence PCR, Shanghai Zhijiang, China, NO. P20200203) were used for nucleic acid detection. This kit uses one step RT-PCR combined with Taqman technology to detect RdRp, E and N genes. Positive was concluded if RdRp gene was positive (Ct < 43), and one of E or N was positive (Ct <43). Patients were also diagnosed as positive if two sequential tests of RdRp were positive while E and N were negative. According to the abovementioned guideline, COVID-19 patients are classified into four subgroups: 1) Mild: mild symptoms without pneumonia; 2) Typical: fever or respiratory tract symptoms with pneumonia; 3) Severe: fulfill any of the three criteria: respiratory distress, respiratory rate ≥ 30 times/min; means oxygen saturation ≤ 93% in resting state; arterial blood oxygen partial pressure/oxygen concentration ≤ 300 mmHg (1 mmHg = 0.133 kPa); 4) Critical: fulfill any of the three criteria: respiratory failure and require mechanical ventilation; shock incidence; admission to ICU with other organ failure. In this study, we included both severe and no-severe patients, with the latter composed of mild and typical COVID-19 patients. Last follow-up of these patients was March 10, 2020. We also procured 25 non-COVID-19 patients with similar clinical characteristics including fever and/or cough as COVID-19 patients however negative in the nucleic acid test. Based on the Chinese Government Diagnosis and Treatment Guideline (Trial 5th version) (Medicine, 2020), patients are defined as suspected COVID-19 cases when they meet the following three clinical criteria: 1) fever or respiratory symptoms, 2) imaging manifestation of pneumonia, and 3) optional reduction of white blood cell or lymphocyte count at early stage. The patients only need to meet at least two of the above three criteria if they have been exposed to COVID-19 individuals. We also collected serum samples from 28 healthy individuals as control. All the serum samples were collected as venous whole blood in the early morning before diet using serum separation tubes (Zhejiang GongDong Medical Technology Co., Ltd, China). The blood samples were centrifuged at 3,500 rpm for 10 min for serum collection. The serum samples were frozen at -80°C. The samples from this study is from a clinical trial that our team initiated and registered in the Chinese Clinical Trial Registry with a ID of ChiCTR2000031365. This study has been approved by the Ethical/Institutional Review Board of Taizhou Public Health Medical Center and Westlake University. Contents from patients were waived by the boards.

### Proteomic analysis

Serum samples were inactivated and sterilized at 56°C for 30 min, and processed as previously with some modifications. Five μL serum from each specimen was lysed in 50 μL lysis buffer (8 M urea in 100 mM triethylammonium bicarbonate, TEAB) at 32°C for 30 min. The lysates were reductive with 10 mM tris (2-carboxyethyl) phosphine (TCEP) for 30 min at 32°C, then alkylated for 45 min with 40 mM iodoacetamide (IAA) in darkness at room temperature (25°C). The protein extracts were diluted with 200 μL 100 mM TEAB, and digested with double-step trypsinization (Hualishi Tech. Ltd, Beijing, China), each step with an enzyme-to-substrate ratio of 1:20, at 32°C for 60 min. The reaction was stopped by adding 30 µL 10% trifluoroacetic acid (TFA) in volume. Digested peptides were cleaned-up with SOLAμ (Thermo Fisher Scientific™, San Jose, USA) following the manufacturer’s instructions, and lableled with TMTpro 16plex label reagents (Thermo Fisher Scientific™, San Jose, USA) as described previously. The TMT samples were fractionated using a nanoflow DIONEX UltiMate 3000 RSLCnano System (Thermo Fisher Scientific™, San Jose, USA) with an XBridge Peptide BEH C18 column (300 Å, 5 μm × 4.6 mm × 250 mm) (Waters, Milford, MA, USA)(Gao et al., 2020). The samples were separated using a gradient from 5% to 35% acetonitrile (ACN) in 10 mM ammonia (pH=10.0) at a flow rate of 1 mL/min. Peptides were separated into 120 fractions, which were consolidated into 40 fractions. The fractions were subsequently dried and re-dissolved in 2% ACN/0.1% formic acid (FA). The re-dissolved peptides were analyzed by LC-MS/MS with the same LC system coupled to a Q Exactive HF-X hybrid Quadrupole-Orbitrap (Thermo Fisher Scientific™, San Jose, USA) in data dependent acquisition (DDA) mode. For each acquisition, peptides were loaded onto a precolumn (3 µm, 100 Å, 20 mm*75 µm i.d.) at a flowrate of 6 μL/min for 4 min and then injected using a 35 min LC gradient (from 5% to 28% buffer B) at a flowrate of 300 nL/min (analytical column, 1.9 µm, 120 Å, 150 mm*75 µm i.d.). Buffer A was 2%ACN, 98% H2O containing 0.1% FA, and buffer B was 98% ACN, 2% H2O containing 0.1% FA. All reagents were MS grade. The *m/z* range of MS1 was 350-1,800 with the resolution at 60,000 (at 200 *m/z*), AGC target of 3e6, and maximum ion injection time (max IT) of 50 ms. Top 15 precursors were selected for MS/MS experiment, with a resolution at 45,000 (at 200 *m/z*), AGC target of 2e5, and max IT of 120 ms. The isolation window of selected precursor was 0.7 *m/z*. The resultant mass spectrometric data were analyzed using Proteome Discoverer (Version 2.4.1.15, Thermo Fisher Scientific) using a protein database composed of the *Homo sapiens* fasta database downloaded from UniprotKB on 07 Jan 2020 and the SARS-CoV-2 virus fasta downloaded from NCBI (version NC_045512.2). Enzyme was set to trypsin with two missed cleavage tolerance. Static modifications were set to carbamidomethylation (+57.021464) of cysteine, TMTpro (+304.207145) of lysine residues and peptides’ N termini, and variable modifications were set to oxidation (+15.994915) of methionine and acetylation (+42.010565) of peptides’ N-termini. Precursor ion mass tolerance was set to 20 ppm, and product ion mass tolerance was set to 0.06 Da. The peptide-spectrum-match allowed 1% target false discovery rate (FDR) (strict) and 5% target FDR (relaxed). Normalization was performed against the total peptide amount. The other parameters followed the default setup.

### Quality control of proteomic data

The quality of proteomic data was ensured at multiple levels. First, a mouse liver digest was used for instrument performance evaluation. We also run water samples (buffer A) as blanks every 4 injections to avoid carry-over. Serum samples of four patient groups from both training and validation cohorts were randomly distributed in eight different batches. Six samples were injected in technical replicates.

### Metabolomic analysis

Ethanol was added to the serum samples and shaken vigorously to inactivate any potential viruses, then dried in a biosafety hood. The dried samples were further treated for metabolomics analysis. The metabolomic analysis was performed as described previously(Lee et al., 2019). Briefly, deactivated serum samples, 100 μL each, were extracted by adding 300 μL extraction solution. The mixtures were shaken vigorously for 2 min. Proteins were denatured and precipitated by centrifugation. The supernatants contained metabolites of diverse chemical natures. To ensure the quantity and reliability of metabolite detection, four platforms were performed with non-target metabolomics. Each supernatant was divided into four fractions: two for analysis using two separate reverse-phase /ultra-performance liquid chromatography (RP/UPLC)-MS/MS methods with positive ion-mode electrospray ionization (ESI), one for analysis using RP/UPLC-MS/MS with negative-ion mode ESI, and one for analysis using hydrophilic interaction liquid chromatography (HILIC)/UPLC-MS/MS with negative-ion mode ESI. Each fraction was dried under nitrogen gas to remove the organic solvent and later re-dissolved in four different reconstitution solvents compatible with each of the four UPLC-MS/MS methods.

All UPLC-MS/MS methods used ACQUITY 2D UPLC system (Waters, Milford, MA, USA) and Q Exactive HF hybrid Quadrupole-Orbitrap (Thermo Fisher Scientific™, San Jose, USA) with HESI-II heated ESI source and Orbitrap mass analyzer. The mass spectrometer was operated at 35,000 mass resolution (at 200 *m/z*). In the first UPLC-MS/MS method, the QE was operated under positive electron spray ionization (ESI) coupled with a C18 column (UPLC BEH C18, 2.1 × 100 mm, 1.7 μm; Waters) was used in UPLC. The mobile solutions used in the gradient elution were water and methanol containing 0.05% perfluoropentanoic acid (PFPA) and 0.1% FA. In the second method, the QE was still operated under ESI positive mode, and the UPLC used the same C18 column as in method one, but the mobile phase solutions were optimized for more hydrophobic compounds and contained methanol, acetonitrile, water, 0.05% PFPA, and 0.01% FA. The third method had the QE operated under negative ESI mode, and the UPLC method used a C18 column eluted with mobile solutions containing methanol and water in 6.5 mM ammonium bicarbonate at pH 8. The UPLC column used in the fourth method was HILIC column (UPLC BEH Amide, 2.1 × 150 mm, 1.7 μm; Waters), and the mobile solutions were consisted of water and acetonitrile with 10 mM ammonium formate at pH 10.8; the QE was operated under negative ESI mode. The QE mass spectrometer analysis was alternated between MS and data-dependent MS2 scans using dynamic exclusion. The scan range was 70-1,000 *m/z*.

After raw data pre-processing, peak finding/alignment, and peak annotation using in-house software, metabolites were identified by searching an in-house library containing more than 3,300 standards with library data entries generated from running purified compound standards through the experimental platforms. Identification of metabolites must meet three strict criteria: narrow window retention index (RI), accurate mass with variation less than 10 ppm and MS/MS spectra with high forward and reverse scores based on comparisons of the ions present in the experimental spectrum to the ions present in the library spectrum entries. Almost all isomers can be distinguished by these three criteria. All identified metabolites meet the level 1 requirements by the Chemical Analysis Working Group (CAWG) of the Metabolomics Standards Initiative (MSI) expect some asterisk labeled lipids which MS/MS spectral were *in silico* matched.

### Quality control of metabolomics analysis

Several types of quality control samples were included in the experiment: a pooled sample generated by taking a small volume of each experimental sample to serve as a technical replicate that was run multiple times throughout the experiment, extracted water samples served as blanks, and extracted commercial plasma samples for monitoring instrument variation. A mixture of internal standards was also spiked into every sample to aid chromatographic peak alignment and instrument stability monitoring. Instrument variability was determined by calculating the median relative SD (RSD) of all internal standards in each sample. The experimental process variability was determined by calculating the median RSD for all endogenous metabolites present in the pooled quality control samples.

### Statistical analysis

Metabolites and therapeutic compounds with over 80% missing ratios in a particular patient group were removed for the metabolomics dataset containing endogenoeous metabolites while full proteomics features were used for the subsequent statistical analysis. Missing values were imputated with the minimal value for each feature. Log2 fold-change (log2 FC) was calculated on the mean of the same patient group for each pair of comparing groups. Two-sided unpaired Welch’s t test was performed for each pair of comparing groups and adjusted p values were calculated using Benjamini & Hochberg correction. The statistical significantly changed proteins or metabolites were selected using the criteria of adjust p value less than 0.05 indicated and absolute log2 FC larger than 0.25. From the training cohort, the important features were selected with mean decrease accuracy larger than 3 using random forest containing a thousand trees using R package randomForest (version 4.6.14) random forest analysis with 10-fold cross validation as binary classification of paired severe and non-severe group using combined differentially regulated proteins and metabolites features. The random forest analysis was further performed for a hundred times on the matrix with only the selected important features using normalized additive predicting probability as the final predicting probability and the larger probability as the predictive label. Those selected important features were used for the random forest analysis on the independent validation cohort.

### Pathway analysis

Four network pathway analysis tools were used for pathway analysis using 93 differentially expressed proteins (DEPs). The top Gene Ontology (GO) processes were enriched by Metascape web-based platform (Zhou et al., 2019). The GO terms is enriched using the Cytoscape plug-in ClueGO (Bindea et al., 2009). Ingenuine pathway analysis (Kramer et al., 2014) of the regulated proteins identifies most significantly relevant pathways with p value of determined based on right-tailed Fisher’s Exact Test with the overall activation or inhibition states of enriched pathways were predicted by z-score. Functional co-expression network analysis by GeNet(Li et al., 2018a) to represent statistical co-expressed protein modules.

**Figure S1.**
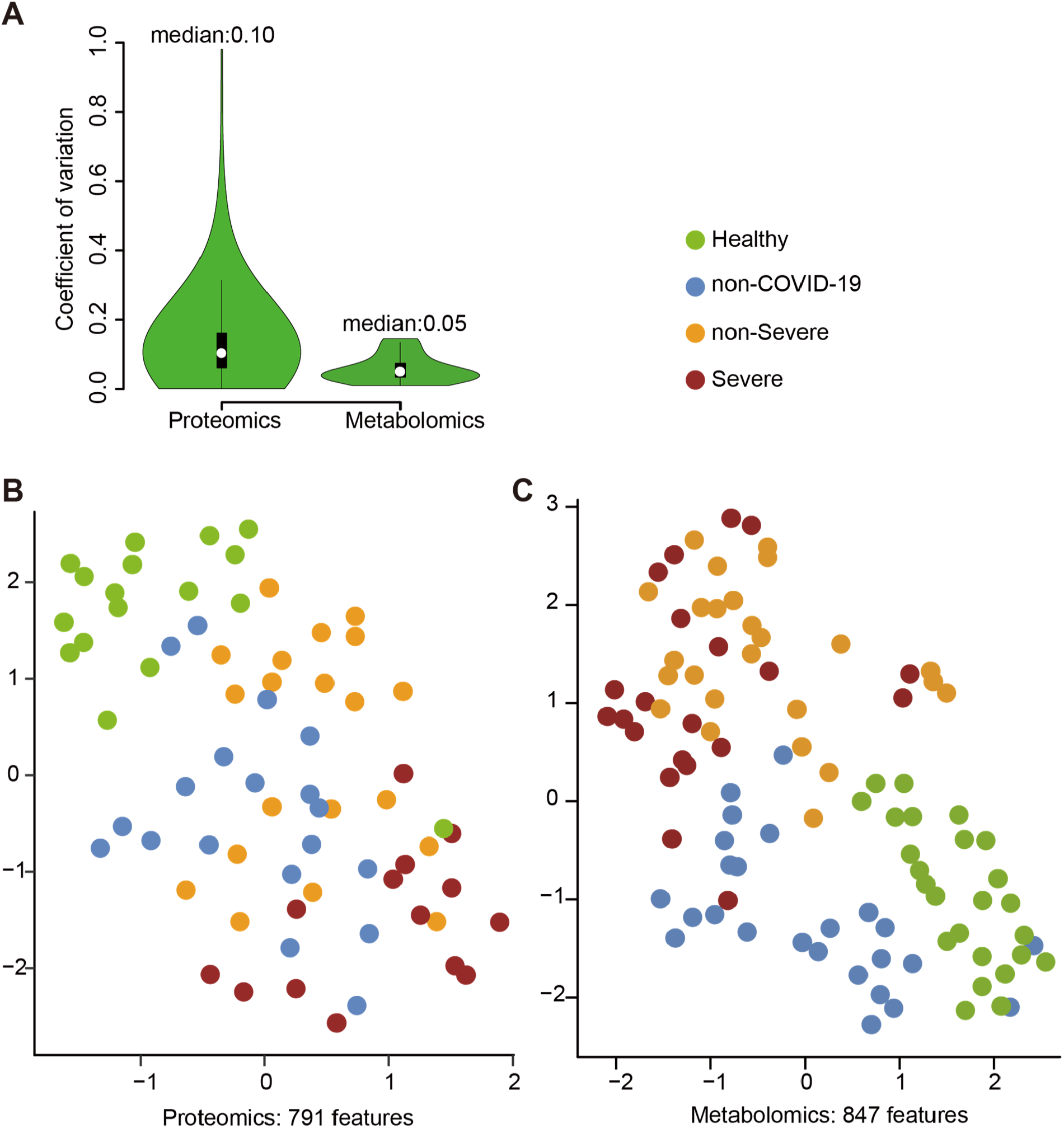
Quality control of proteomic and metabolomic data. (A) Coefficient of variation (CV) of the proteomic data is calculated by the proteins quantified in six quality control (QC) samples using the pooled samples from all samples in training cohort. CV of the metabolomic data is calculated by twelve QC samples using a set of isotopic internal spiked-in standards. (B) UMAP of sera samples using 791 measured proteins. (C) UMAP of sera samples using 847 metabolites.

**Figure S2.**
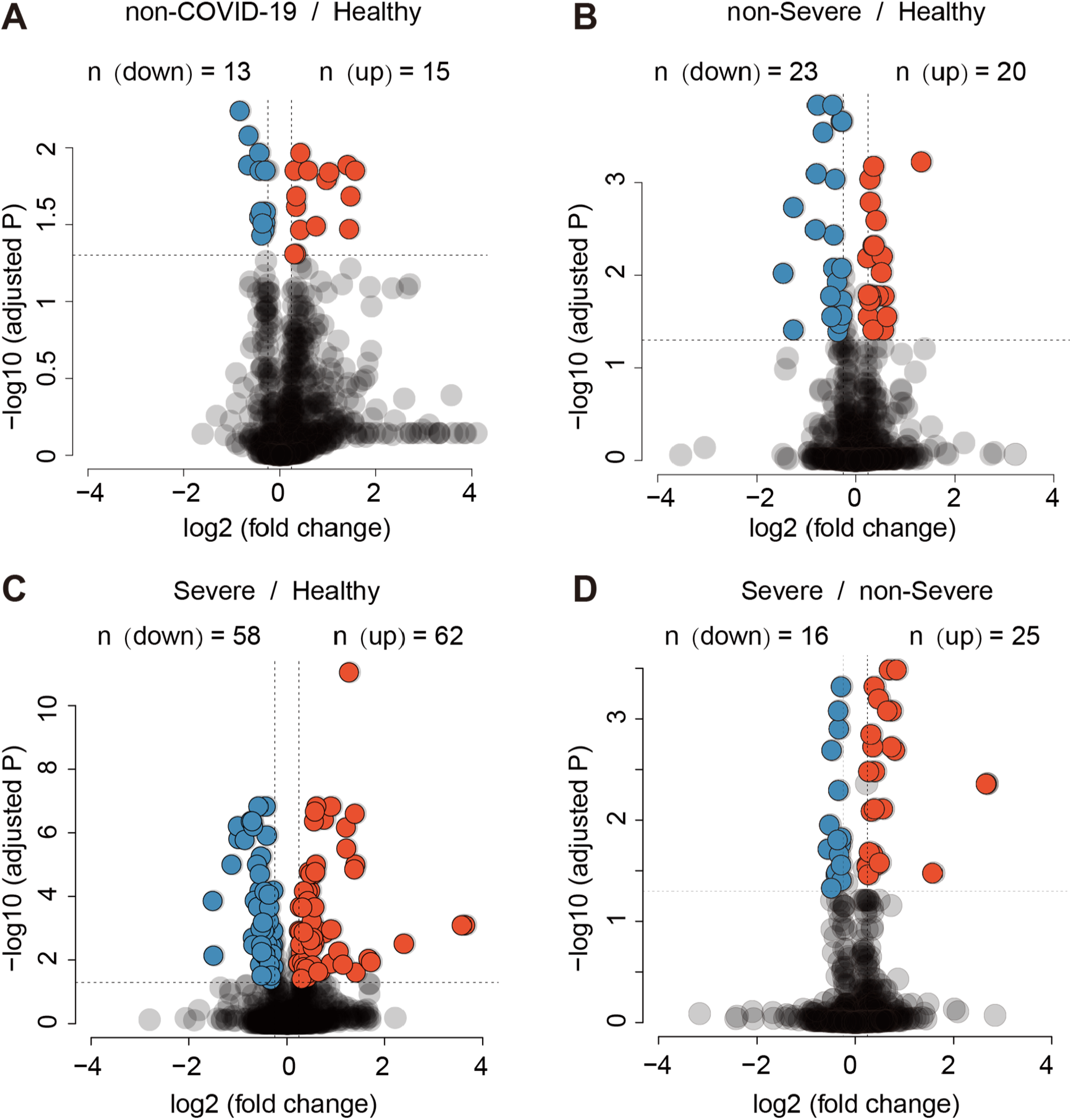
Differentially expressed proteins in different patient groups in the training cohort. Volcano plots compare four pairs of patient groups as indicated in the plot. Proteins with log2 (fold-change) beyond 0.25 or below - 0.25 with adjusted p value lower than 0.05 were considered as significantly differential expression. Number of significantly down- (blue) and up- (red) regulated proteins were shown on the top.

**Figure S3.**
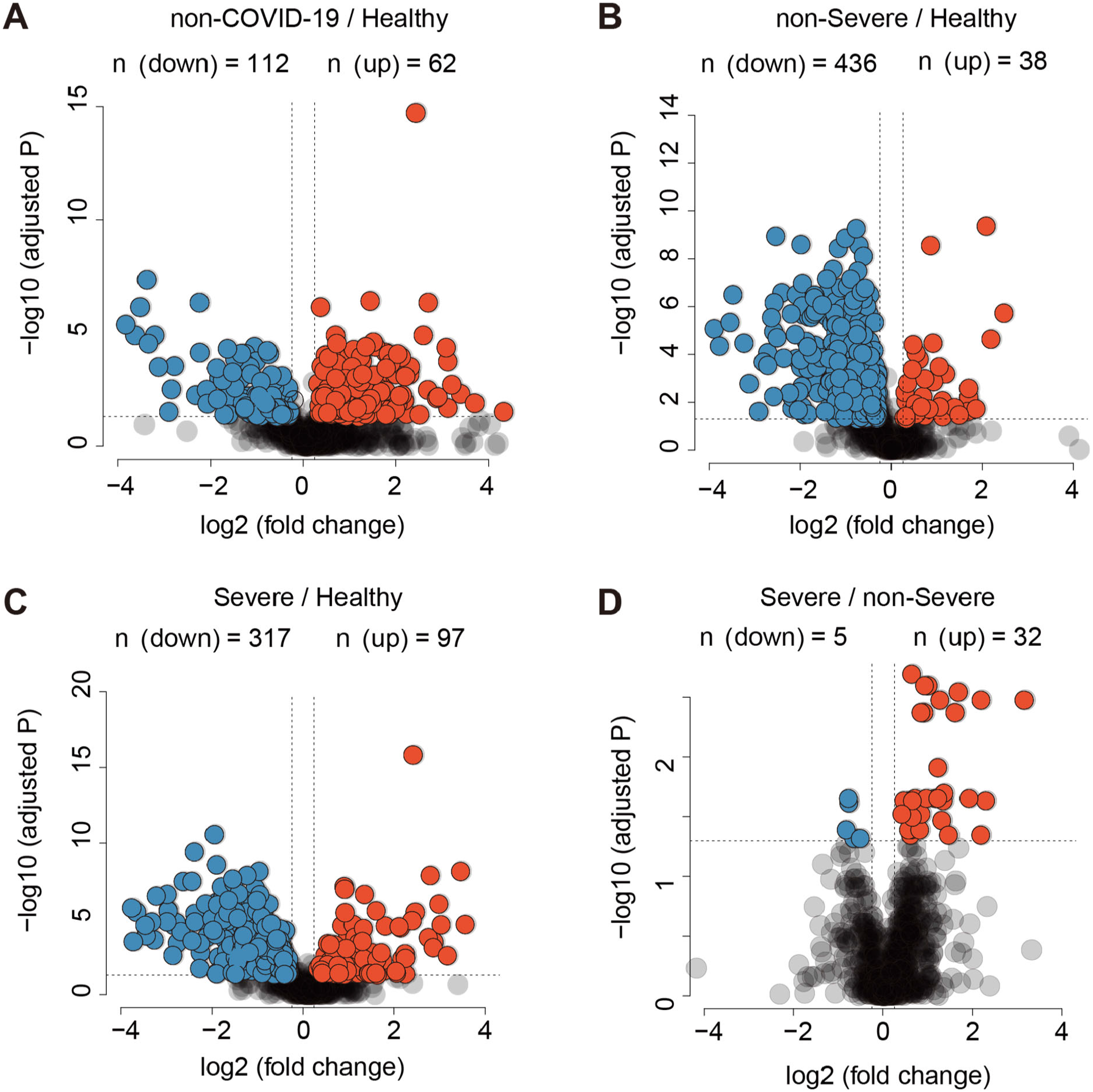
Differentially expressed metabolites in different patient groups in the training cohort. The volcano plots follow that for Figure S3 except metabolomic data were used as the input.

**Figure S4.**
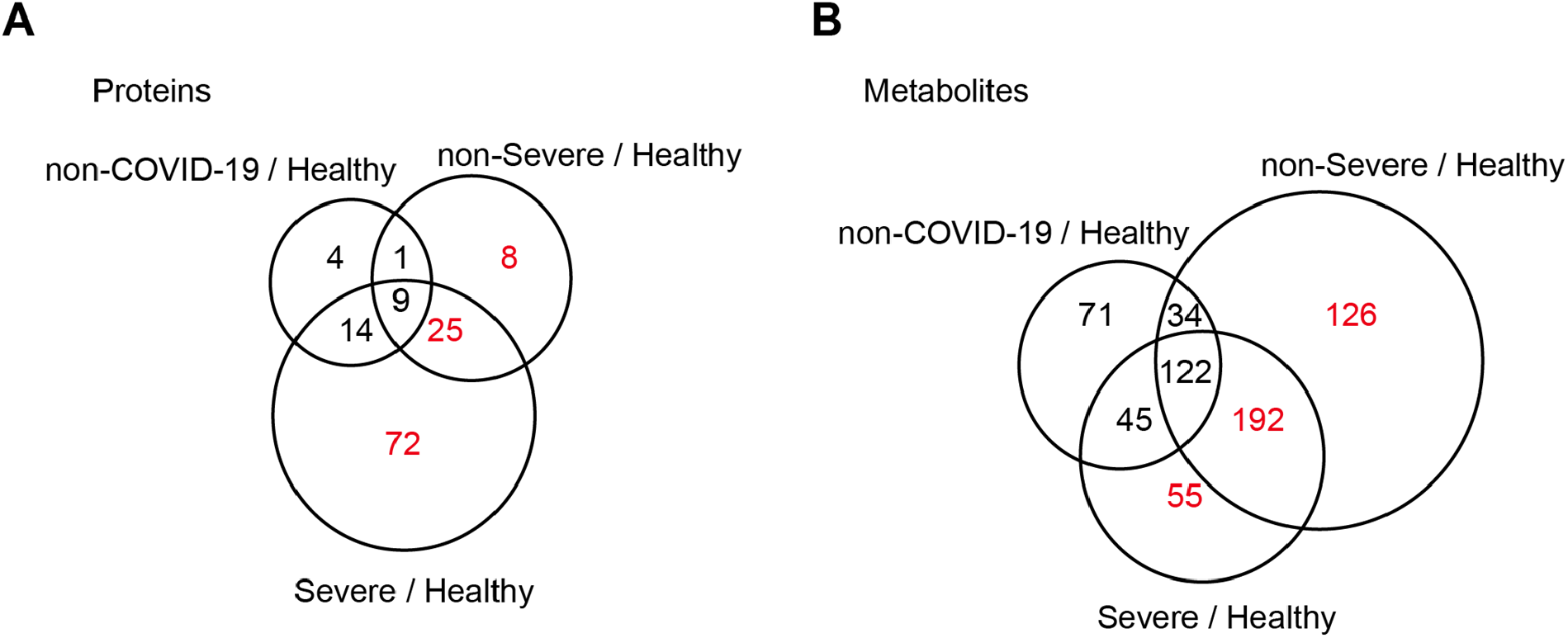
Proteins and metabolites regulated in COVID-19 patients but not in non-COVID-19 patients. Venn diagrams showing the overlaps between significantly regulated proteins (A) and metabolites (B) as identified in volcano plots. Proteins and metabolites labeled in red are the shortlisted molecules which differentially expressed in the COVID-19 patients but not in the non-COVID-19 patients.

**Figure S5.**
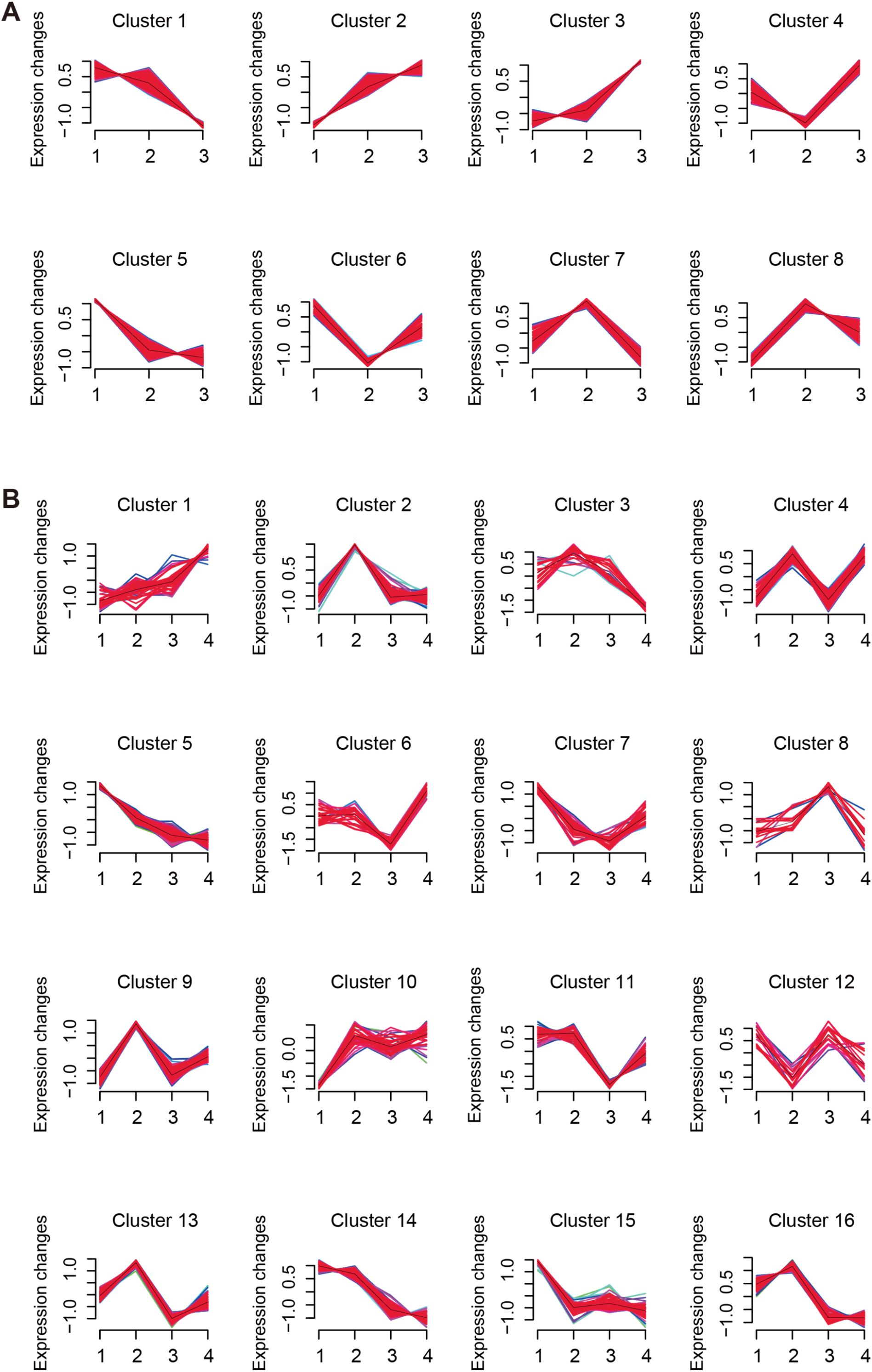
Identification of specific clusters of proteins and metabolites in COVID-19 patients. 791 proteins (A) and 941 metabolites (B) were clustered using mFuzz into 16 significant discrete clusters, respectively, to illustrate the relative expression changes of the proteomics and metabolomics data. The groups in proteomics data: 1: Healthy; 2: non-Severe COVID-19; 3: Severe COVID-19. The groups in metabolomics data: 1: Healthy; 2: non-COVID-19; 3: non-Severe COVID-19; 4: Severe COVID-19.

**Figure S6.**
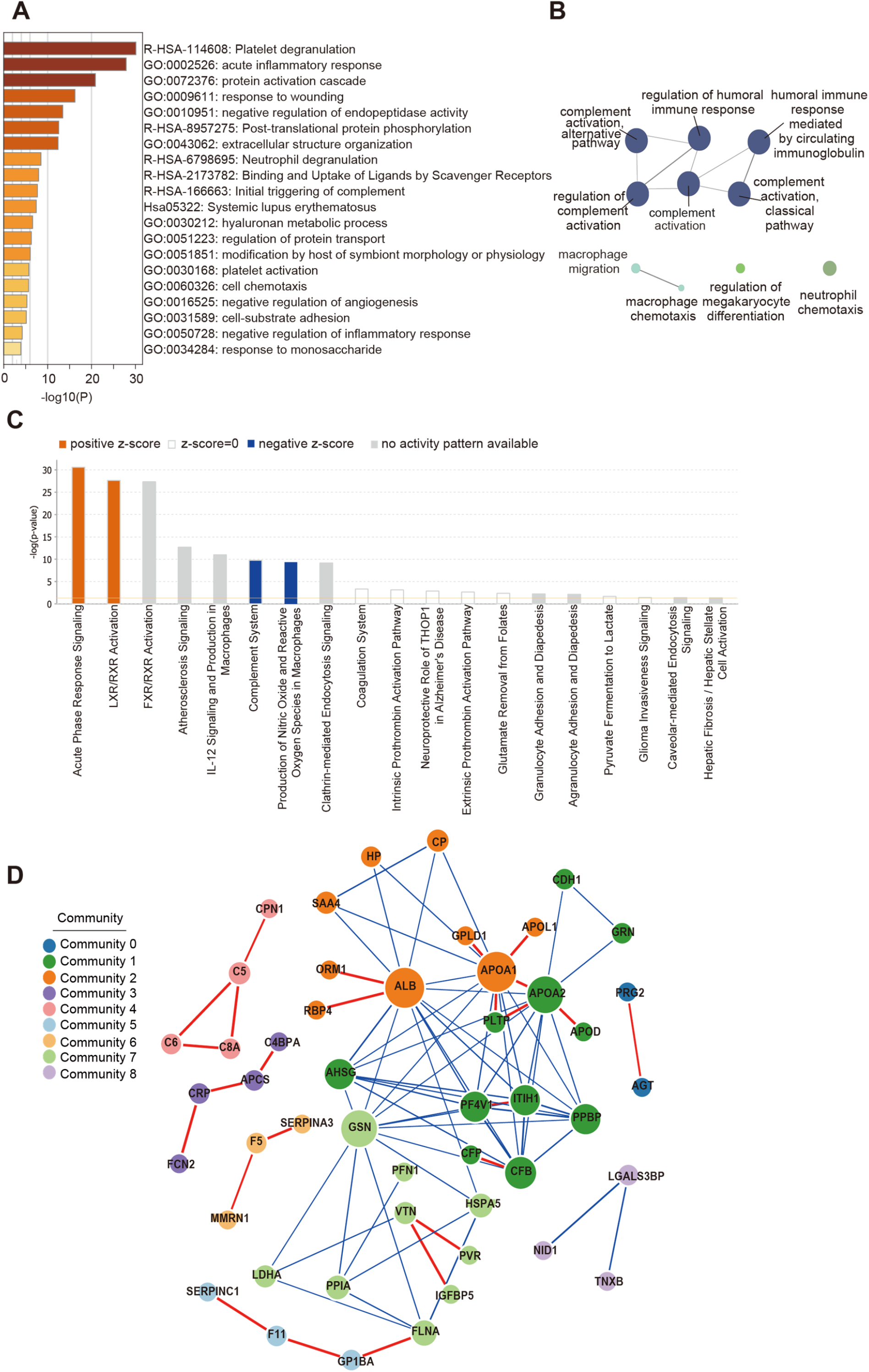
Pathway analysis of 93 differential expressed proteins in COVID-19 patients. (A) The Gene Ontology (GO) processes enriched by Metascape. (B) The GO terms enriched using the Cytoscape plug-in ClueGO. Ingenuine pathway analysis of most significantly relevant pathways with the predicted activation or inhibition state. (D) Functional network analysis by GeNet identifies several communities.

